# Cost-effectiveness of a 4 days-a-week triple therapy in persons living with HIV: an ancillary study of the ANRS 170 QUATUOR noninferiority trial

**DOI:** 10.1101/2024.09.26.24314433

**Authors:** Gilles Hejblum, Samih Daher, Paul Moulaire, Karine Amat, Sidonie Lambert-Niclot, Clotilde Allavena, Christine Katlama, Karine Lacombe, Diane Ponscarme, Jade Ghosn, Severine Gibowski, Jean-Claude Alvarez, Jacqueline Capeau, Laurence Morand-Joubert, Dominique Costagliola, Pierre De Truchis, Roland Landman, Lambert Assoumou

**Author notes:** **Corresponding author :** Gilles Hejblum, Institut Pierre Louis d’Épidémiologie et de Santé Publique, 27 rue Chaligny, 75012 Paris, France.

## Abstract

**Background:** ANRS 170 QUATUOR study (NCT03256422) demonstrated the noninferiority of a triple antiretroviral therapy (ART) taken 4 days a week (4/7-days) compared to a triple therapy taken 7 days a week (7/7-days) for persons living with HIV and with suppressed viremia. We aimed at investigating corresponding cost-effectiveness issues.

**Methods:** All persons involved in the primary analysis of the QUATUOR noninferiority trial were considered (N=636, 318 per arm) in this cost-effectiveness study. Time horizon was the first 48 weeks of the trial. Effectiveness was considered as the noninferiority of the 4/7-days strategy, main criterion of the trial primary analysis. Direct health resource costs (_year2022_€) were considered and included costs for ART, laboratory tests, co-medications, hospitalizations, and medical consultations. Analyses were based on 10,000 simulations replicating the trial. Additional analyses explored the national impact of spreading the 4/7-days strategy all over France. Sensitivity analyses included considering treatment success as effectiveness, and various proportions of persons adopting the 4/7-days strategy.

**Findings:** The mean individual total costs amounted to € 5,049 [95% confidence interval: 4,798–5,345] and € 8,089 [7,706–8,527] in the 4/7-days and 7/7-days arm, respectively. Corresponding ART costs were € 3,678 [3,593–3,763] and € 6,450 [6,301–6,596], respectively (p<0.0001), and constituted the single cost element with a statistical difference between the two strategies. Considering noninferiority as the effectiveness criterion in a health economic perspective, the 4/7-day strategy provided average savings of € 3,013 [2829–3196] in ART costs per individual and per year. Adopting the 4/7-days regimen in 20% of the potential recipients in France was estimated to provide annual savings of € 61,752,467 [61,569,005; 61,925,136].

**Interpretation:** The 4/7-day strategy dominates the 7/7-day strategy since it spares substantial ART costs while maintaining treatment effectiveness. Study results support generalizing the adoption of 4/7-days triple ART regimens in France, when possible.

## INTRODUCTION

According to international guidelines, antiretroviral therapy (ART) for persons living with HIV (PLWH), whether naive or experienced, is based on a regimen of three or two drugs taken seven days a week.^1^ However, as treatment lasts a lifetime, the cumulative exposure to such drugs raises several concerns relating to long-term drug toxicity.^2^ Therefore, several strategies have been investigated for reducing maintenance therapy in patients with suppressed viremia, including intermittent maintenance treatment in short cycles. Leibowitch’s retrospective study showed that a triple ART taken 4 consecutive days on and 3 days off (further referred to as a 4/7-days strategy) resulted in maintained viral suppression.^3^ These results led to a multicenter prospective pilot study in participants receiving a triple ART based on a non-nucleoside reverse transcriptase inhibitor or a boosted protease inhibitor, with a therapeutic success rate of 96% after 48 weeks.^4^ The ANRS-170 QUATUOR trial was conducted to confirm the efficacy of a 4/7-days strategy in terms of virological success, safety, adherence and quality of life.^5^ This randomized noninferiority trial demonstrated that over a 48-week period, the intermittent short-cycle 4/7-days strategy was noninferior to a standard daily treatment (further referred to as a 7/7-days strategy) with the most widely used treatment regimens, including integrase inhibitors and nonnucleoside reverse transcriptase inhibitors.

The present cost-effectiveness study compares the 7/7-days and the 4/7-days strategies, based on detailed investigations on QUATUOR data relating to health economic aspects. In addition, we estimated the annual savings in France that would potentially occur if the 4/7 strategy was deployed throughout the whole country in eligible PLWH.

## METHODS

### Brief description of the QUATUOR trial and corresponding data collection

Participants were assessed at screening, baseline, and weeks 4, 12, 24, 36, and 48. At each visit, participants had routine safety monitoring and collection of blood samples for full blood cell counts, serum chemistry, liver, renal function and plasma viral load measurements. CD4+ and CD8+ cell counts were measured at screening, week 12, week 24, and week 48. Fasting serum lipids were measured at baseline, week 24 and week 48. Any plasma viral load value >50 copies/mL during follow-up was confirmed on a new sample taken 2-4 weeks later. If the value at control was >50 copies/mL, the participant was considered to be in virological failure, and drug resistance genotyping and drug assays were performed. Participants randomized to the 4/7-days strategy took the ART for 4 consecutive days and stopped it for the following 3 days, and so on for 48 weeks. Participants randomized to the 7/7-days strategy took ART daily until week 48. ART was prescribed at baseline, then every 12 weeks until week 48. Any change in the ART medication or strategy during the 48-week of follow-up was recorded including the date of change. All co-medications prescribed during the 48 weeks of follow-up were also recorded in the electronic case report form. Serious adverse events were reported in electronic case report form. In the case of hospitalization, the date of hospitalization and the date of discharge were collected.

### Type of analysis and population studied

The cost-effectiveness analysis of a randomized noninferiority trial is reported here according to CHEERS international guidelines,^6^ and was conducted according the French recommendations.^7^ The cost-effectiveness analysis compared the 4/7-days and the 7/7-days strategies. The population studied was the 636 PLWH that were analyzed in the main analysis of the QUATUOR trial,^5^ 318 individuals in each strategy arm.

### Perspective and time horizon of the study

Aiming at contributing to recommendations of health authorities, the study perspective was that of the mandatory health insurance in France. Time horizon was that of the QUATUOR main analysis,^5^ i.e., 48 weeks. Accordingly, no discounting rate was applied in the analysis.

### Effectiveness

The main criterion retained for expressing the effectiveness of the 4/7 strategy as compared to the 7/7 strategy was the noninferiority criterion that was adopted in the QUATUOR analysis. We also explored as an additional effectiveness criterion the individual success of the treatment, using FDA Snapshot algorithm.^8^

### Costs

Investigations on costs were based on the official prices in France in year 2022, and are reported in Euros. The costs of the following direct medical consumptions were considered in the study: ART,^9^ biological tests / measures,^10^ hospitalizations,^11^ medical consultations,^12^ and co-medication prescriptions^9^ that were collected at the individual level in the case report forms of the trial. All combinations of ART taken individually were duly documented each week allowing an individual cost for ART consumption. Co-medications, medical consultations, and hospitalizations were also documented at the individual level. Co-medications included prescriptions for the management of adverse events or medications for other reasons, for example chronic diseases, or occasional therapies (e.g., infections such as influenza-like illness). Information on hospitalizations was limited to a short description of the reason for the hospital stay as well as the length of stay. The most likely code of hospital stay was derived from this information, and the cost of the hospital stay was valued according to the price applied by the health care system in France in 2022.^11^

### Cost-effectiveness

As recommended,^6^ the main judgement criterion retained for cost-effectiveness analyses was the incremental cost-effectiveness ratio (ICER): difference of the costs between the two strategies divided by the corresponding difference of effectiveness. As aforementioned, the results of the QUATUOR trial indicated that the noninferiority criterion was reached by the 4/7-days strategy. Therefore, in the main analysis considering the noninferiority criterion as effectiveness, the analysis especially investigated the differences between the 4/7-days and 7/7-days strategies relating to the cost elements.

Considering the analyses in which the individual treatment success was considered as the effectiveness criterion, the incremental cost-effectiveness ratio corresponds to the additional cost required by a strategy (as compared to the alternative strategy) for enabling an additional treatment success.

### Statistical analyses

Analyses were made with R statistical software, version 4.2.2. The 95% confidence intervals (CI) reported were all obtained by bootstrap. The statistical significance of the differences observed between the 4/7-days and 7/7-days strategies was assessed with permutation tests,^13^ based on 10,000 replications of the QUATUOR trial in which the 636 individuals were randomly assigned to one of the arms (318 in each arm) for determining the distribution of the differences under the null hypothesis. Cost-effectiveness analyses considering individual treatment success as the effectiveness criterion were based on 10,000 simulations of the QUATUOR trial. In each of these simulations, 318 individuals in each arm were simulated (same sample size as the original trial), the corresponding individuals being randomly chosen in the initial pool of the 318 individuals in the arm of the effective QUATUOR trial, but allowing the resampling of individuals.

### Extending analyses to predicting the economic impact of the 4/7-days strategy at the national French level

Based on the cost-effectiveness estimates obtained with QUATUOR data, we developed extended analyses estimating the economic impact of applying the 4/7-days strategy at a national level. Such an impact was estimated considering three time horizons, 1, 5 and 10 years. Undiscounted estimates were considered according to French^14^ and international^15^ recommendations for reporting studies on budget impact. Sensitivity analyses reporting discounted estimates were additionally conducted, and two discounting rates considered: a rate of 4% was applied according to French recommendations for studies investigating model-based decisions in health economics,^7^ and a rate of 3% was additionally considered as a sensitivity analysis. The study assumed that the persons potentially eligible for adopting a 4/7-days strategy in France were the 104,042 individuals enrolled in the French Hospital Database on HIV (ANRS CO4-FHDH), who initiated ART before 2017 and had at least one follow-up visit between 1 January 2017 and 31 December 2019, as reported by Marty et al.^16^ Moreover, the main analysis assumed that in practice, 20% (n = 20,808) or 50% (n = 52,021) of these potentially eligible individuals would effectively switch from a 7/7-days to a 4/7-days strategy. The age distribution of the 104,042 individuals reported by Marty et al^16^ was used for determining the potential deaths occurring during the 10-year time horizon of the analyses. Death rates according to age and sex in the general population of France were obtained from data publicly available from the French National Institute of Statistics and Economic Studies (Insee).^17-19^ These death rates were considered as baseline death rates to which some excess mortality relating to HIV was added. According to Edwards et al, when considering a period ranging from 1999 to 2017, the excess mortality of persons living with HIV was 7.9%, and this rate declined to 2.6% when restricting this period to years 2011–2017.^20^ Therefore, the present analyses considered that individuals of the cohort had an excess mortality (as compared to the French general population) that linearly decreased each year: 8% the 1^st^ year, (8% x 8/9) the 2^nd^ year, (8% x 7/9) the 3^rd^ year, …, and 0% the 10^th^ year. Sensitivity analyses additionally considered a fixed 8% excess mortality all over the 10-year time horizon, and also no excess mortality at all (i.e., mortality rates in the cohort similar to those of the general population). In practice, the cohort of the above-mentioned 104,042 individuals was replicated in 1000 simulations. In each simulation, each individual was randomly assigned one of the ART given in individuals of the same age in the QUATUOR trial, and a Monte-Carlo process determined individual survival during a 10-year follow-up. Corresponding ART costs were calculated for each individual. Cost estimates reported were calculated as the mean from the 1000 simulations.

## RESULTS

Table 1 shows the direct costs of healthcare resources in the QUATUOR trial. Costs of ART represented the major part of the total direct healthcare costs, with 73% and 80% of these costs attributed to costs for ART in the 4/7-days arm (100 x € 1,169,471 / € 1,605,307) and in the arm 7/7 arm (100 x € 2,051,004 / € 2,572,071), respectively. Moreover, during the 48 weeks of the trial, no difference between the two arms was detected considering the costs for laboratory tests (p = 0.65), co-medications (p = 0.15), hospitalizations (p = 0.20), and medical consultations (p = 0.75), while unsurprisingly, the costs for ART were dramatically different (p < 10^−4^). Therefore, costs for ART constitute the single cost element eventually required in cost-effectiveness analyses comparing the two arms. Noninferiority of the 4/7-days strategy was reached in 7,025 of the 10,000 (70%) simulations that replicated the QUATUOR trial, confirming the main results of the initial analysis of the trial.^5^ Therefore, adopting this noninferiority as the effectiveness criterion in a health economic perspective, the 7/7-days strategy is dominated by the 4/7-days strategy since the former strategy costs more than the second strategy while yielding no additional health effectiveness. More precisely, adopting such a perspective, when considering the 48 weeks of treatment in the QUATUOR trial, a 4/7-days strategy was estimated to provide a mean [95% CI] sparing of € 2,772 [2,603; 2,940] per individual for ART costs as compared to a 7/7-days strategy. Normalizing the difference between the two arms observed during 48 weeks to the difference per individual and per year corresponds to a mean estimate of € 3,013 [2829–3196]. The distributions of ART costs in the two arms differed mainly according to a translation factor (Figure 1A), and were nearly confounded when considering the daily costs only on days of effective medication (Figure 2B), indicating that cost differences between the two arms were solely due to the reduction of days with effective medication in the 4/7-days strategy.

**Table 1:** Costs of health resources in the QUATUOR trial during the 48 weeks of the trial.

| Cost component | 4/7-days arm (n = 318) |  | 7/7-days arm (n = 318) |  | p value* | Incremental Difference (7/7-days – 4/7-days-) |  |
| --- | --- | --- | --- | --- | --- | --- | --- |
|  | Cumulative costs (all individuals) | Costs per individual | Cumulative costs (all individuals) | Costs per individual |  | Cumulative costs (all individuals) | Costs per individual |
| 1: ART (€) | 1,169,471 [1,142,407; 1,196,381] | 3,678 [3,593; 3,763] | 2,051,004 [2,003,639; 2,097,261] | 6,450 [6,301; 6,596] | < 10<sup>-4</sup> | 881,534 [827,543; 934,757] | 2,772 [2,603; 2,940] |
| 2: Laboratory tests (€) | 157,430 [155,157; 159,758] | 496 [488; 503] | 156,838 [154,602; 158,864] | 494 [487; 500] | 0.65 | -593 [-3812; 2475] | -2 [-12; 8] |
| 3: Co-medications (€) | 109,163 [74,823; 168,134] | 344 [236; 529] | 152,034 [93,707; 232,058] | 479 [295; 730] | 0.15 | 42,871 [-43,708; 1332,810] | 135 [-138; 421] |
| 4: Hospitalizations (€) | 121,769 [64,867; 189,505] | 383 [204; 596] | 164,922 [96,428; 242,981] | 519 [304; 765] | 0.20 | 43,153 [-54,011; 139,349] | 136 [-170; 439] |
| 5: Medical consultations (€) | 47,475 [47,175; 47,675] | 150 [149; 150] | 47,275 [46,850; 47,600] | 149 [148; 150] | 0.75 | -200 [-700; 250] | -1 [-3; 1] |
| Total (1+2+3+4+5) | 1,605,307 [1,525,613; 1,699,845] | 5,049 [4,798; 5,345] | 2,572,071 [2,450,605; 2,711,695] | 8,089 [7,706; 8,527] | < 10<sup>-4</sup> | 966,765 [812,698; 1,122,253] | 3,041 [2,556; 3,529] |

All cost estimates correspond to the mean [95% confidence intervals] from 10,000 replications of the QUATUOR trial.
\*Difference between arms.
Abbreviations used: ART, anti-retroviral therapy.

**Figure 1.**
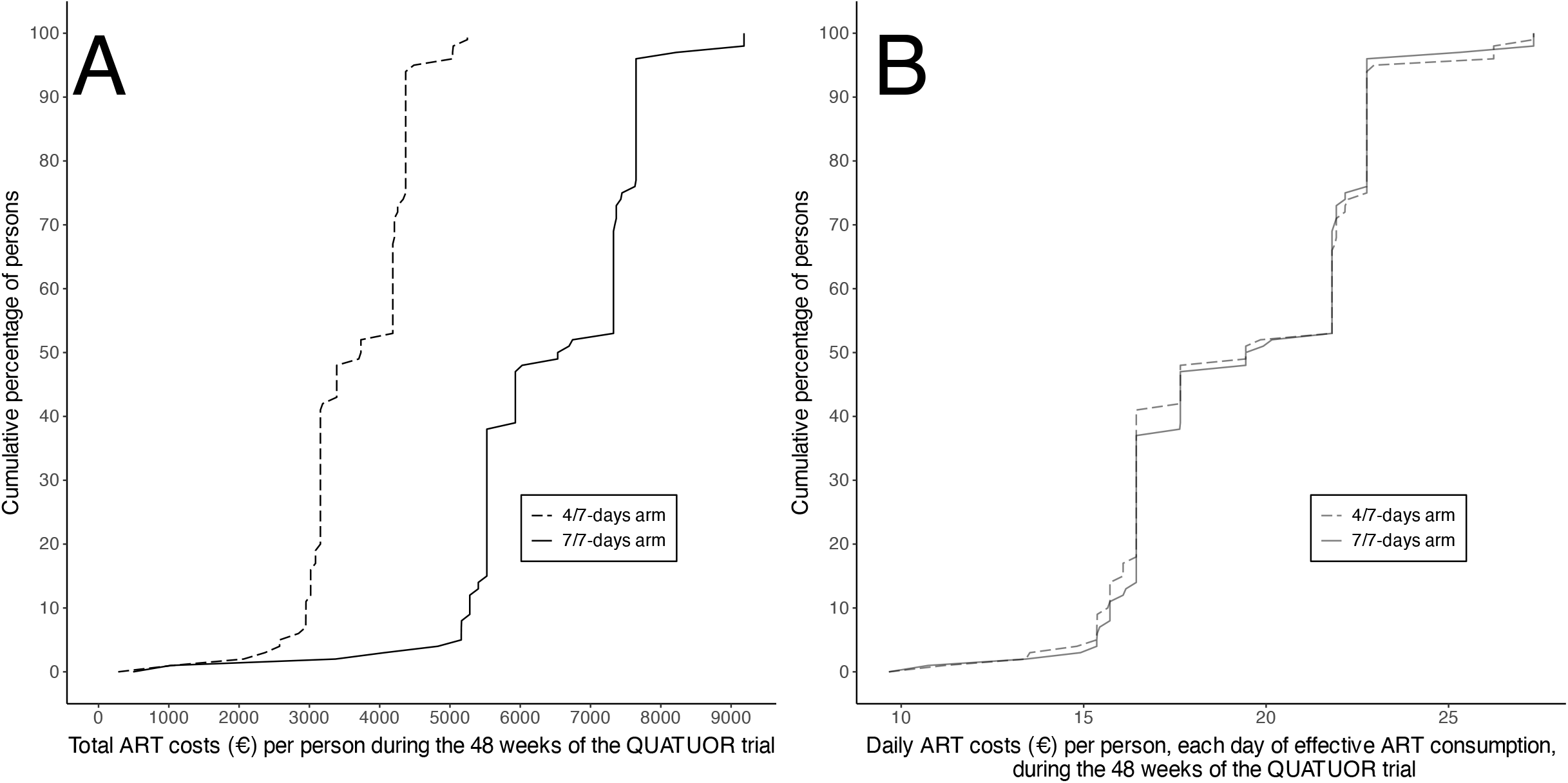
Distribution of the ART costs per person in the group with a 4 days-a-week regimen (4/7-days arm) and in the group with a 7 days-a-week regimen (7/7-days arm).

**Figure 2.**
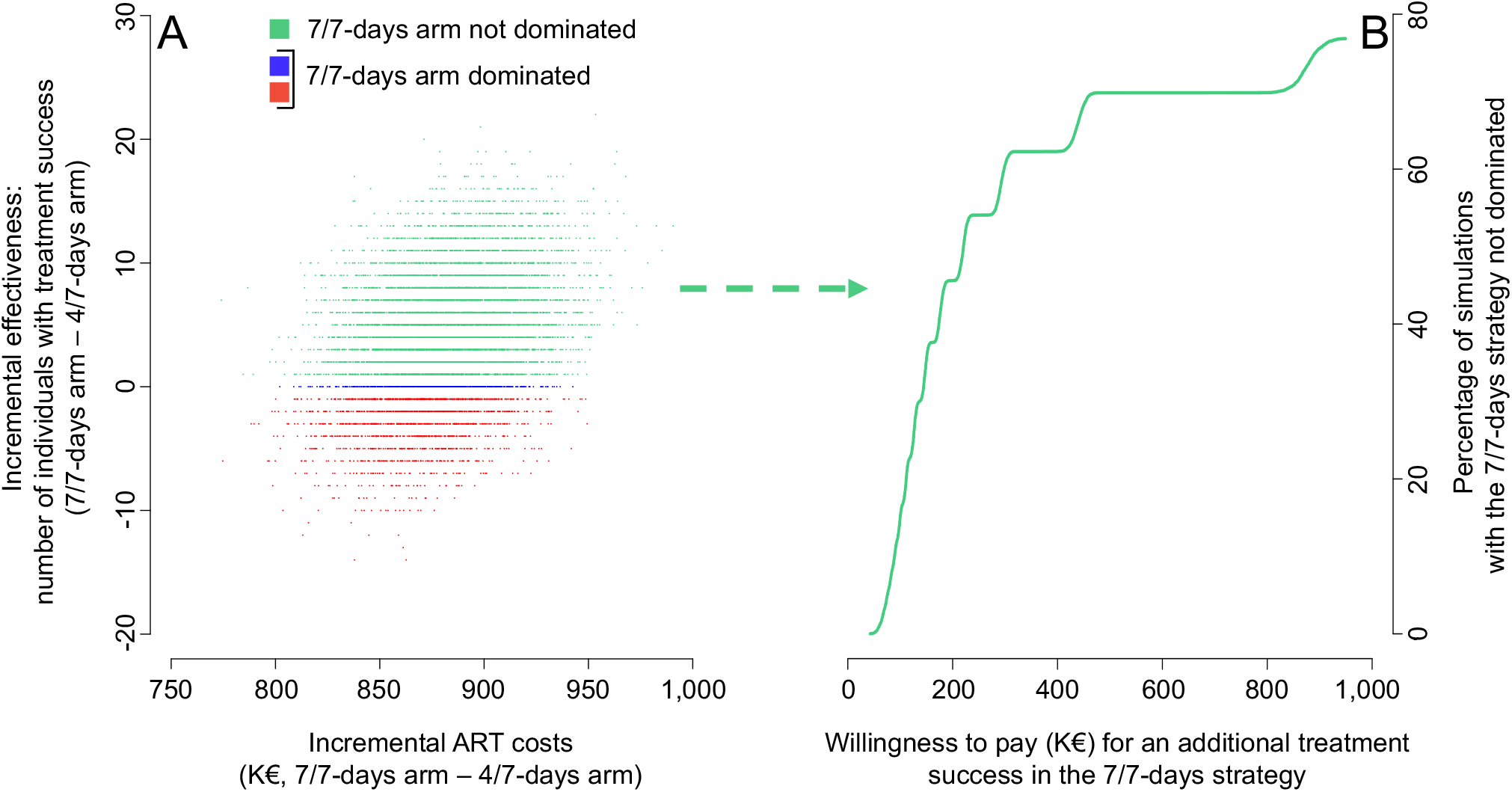
Relationship between ART costs and effectiveness when considering the number of treatment successes observed in the arm (individuals with HIV RNA <50 copies/mL after 48 weeks of treatment without strategy modification) as the effectiveness criterion; results based on 10,000 simulations replicating the QUATUOR trial. Panel A: each dot (n = 10,000) plots the incremental effectiveness and the incremental ART costs issued from one simulation of the trial. Unsurprisingly, all simulations resulted in greater costs observed in the 7/7-days arm. Red dots (n = 1,745) correspond to simulations in which fewer treatment successes were observed in the 7/7-days arm, blue dots (n = 570) correspond to simulations in which identical numbers of treatment success were observed in both arms, and green dots (n = 7,685) correspond to simulations in which more treatment successes were observed in the 7/7-days arm. Panel B: cumulative distribution of the willingness to pay for an additional treatment success, based on the above-mentioned 7,685 simulations in which more treatment successes and greater costs were both observed in the 7/7-days arm.

A sensitivity analysis contrasted the greater costs observed in the 7/7-days arm with the greater proportion of individual treatment successes (308 / 318) that were observed in this arm as compared to the corresponding proportion observed in the 4/7-days arm (304 / 318): adopting treatment success as the effectiveness criterion, the ICER of 7/7-days versus 4/7-days would be € 881,534€ / (308 - 304) = € 220,383 per additional success. In other words, a 7/7-days regimen requires additional ART costs of € 220,383 for observing an additional individual success, as compared to a 4/7-days regimen.

The replicated simulations of the QUATUOR trial provide additional insights on cost-effectiveness issues:

First, the 7/7-days strategy was dominated by the 4/7-days strategy in 2,315 (23%) simulations (Figure 2A, red and blue dots): while ART costs in the 7/7-days arm were greater than those in 4/7-days arm in the corresponding simulations (actually, ART costs were unsurprisingly greater in the 7/7-days arm than in the 4/7-days arm in each of the 10,000 simulations made), there were 1,745 simulations with fewer individual successes observed in the 7/7-days arm than in the 4/7-days arm (Figure 2A, red dots), and 570 additional simulations in which identical numbers of individual successes were observed in both arms (Figure 2A, blue dots).

Second, considering the remaining 7,685 (77%) simulations in which the 7/7-days arm was not dominated by the 4/7-days arm (i.e., greater costs and greater number of successes both observed in the7/7-days arm, green dots in Figure 2A), the ICER of the 7/7-days arm versus 4/7-days ranged from € 42,775 to € 949,164 per additional treatment success, with 1,514 (15%), 4,559 (46%), 6,053 (61%), 6,227 (62%) and 6,958 (70%) simulations with an ICER lower than K€ 100, K€ 200, K€ 300, K€ 400, and K€ 500, respectively (Figure 2B); there was not any simulation with an ICER below 1 gross domestic product per inhabitant (€ 39,323), and there were 2,265 (23%) simulations with an ICER lower than 3 gross domestic product per inhabitant (€ 117,969).

In the end, the replicated simulations of the QUATUOR trial estimated that the mean difference of the number of successes observed in the two arms was not significant (mean [95%CI] = 4[-5; 13]) and the permutation test confirmed that the proportions of successes observed in the two arms of the trial were not significantly different (p = 0.15).

The predicted economic impact of generalizing a switch from a 7/7-days to a 4/7-days strategy in 20% of persons eligible for such a switch in France (n = 0.2 × 104,042 = 20,808) was estimated (Table2). the corresponding total ART costs spared would amount to € 61,752,467 [61,569,005; 61,925,136] the first year (upper part of Table 2). If the time horizon of the prediction was extended to 10 years, the corresponding savings would totalize € 598,179,732 [596,224,428; 600,155,415]. These estimates were obtained assuming an excess mortality decreasing with time each year (from 8% to 0%) in the target population as compared to the general population in France, and sensitivity analyses in which excess mortality was varied (from a fixed excess mortality of 8% each year to no excess mortality at all, see middle and bottom parts of Table 2) indicated that such a variability resulted in very small changes in the estimates in terms of life years per individual, and economic savings. Analyses hypothesizing a scenario more favorable to the adoption of the 4/7-days strategy, with 50% of the eligible persons switching to a 4/7-days strategy, estimated annual savings of € 154,376,338 [154,084,190; 154,659,273] at the national level, and savings of € 1,495,428,616 [1,492,194,706; 1,498,663,767] considering a 10-year time horizon (Table 3).

**Table 2.** Prediction of the total ART costs spared (€) hypothesizing that 20% of the persons living in France and fulfilling QUATUOR inclusion criteria (n = 0.2 × 104,042 = 20,808) would switch from the 7/7-days strategy to the 4/7-days strategy.

| Excess mortality in the target population as compared to the mortality in the whole French population | Time Horizon and discounting rate | Life-years expected for individuals of the target population adopting the 4/7-days strategy |  | ART costs (€) expected to be spared with the 4/7-days strategy as compared to the 7/7-days strategy |
| --- | --- | --- | --- | --- |
| | | Totalized for the whole target population ($n = 20,808$) | per individual | Totalized for the whole target population ($n = 20,808$) |
| Linear decreasing of excess mortality each year (from 8% to 0%)* | 1 year, undiscounted | 20,750 [20,737; 20,761]** | 0.997 | 61,752,467 [61,569,005; 61,925,136]** |
|  | 5 years, undiscounted | 102,465 [102,333; 102,601] | 4.924 | 304,857,074 [303,889,336; 305,771,258] |
|  | 5 years, discounting rate = 3% | 96,704 [96,583; 96,831] | 4.647 | 287,720,062 [286,805,157; 288,580,058] |
|  | 5 years, discounting rate = 4% | 94,928 [94,810; 95,052] | 4.562 | 282,436,923 [281,538,309; 283,280,659] |
|  | 10 years, undiscounted | 201,130 [200,743; 201,509] | 9.666 | 598,179,732 [596,224,428; 600,155,415] |
|  | 10 years, discounting rate = 3% | 177,039 [176,716; 177,362] | 8.508 | 526,552,343 [524,858,049; 528,261,559] |
|  | 10 years, discounting rate = 4% | 170,072 [169,767; 170,377] | 8.173 | 505,836,960 [504,209,209; 507,476,873] |
| Fixed 8% excess mortality | 1 year, undiscounted | 20,750 [20,737; 20,761] | 0.997 | 61,752,467 [61,569,005; 61,925,136] |
|  | 5 years, undiscounted | 102,448 [102,315; 102,587] | 4.923 | 304,808,074 [303,842,724; 305,723,962] |
|  | 5 years, discounting rate = 3% | 96,689 [96,566; 96,818] | 4.647 | 287,675,685 [286,766,982; 288,537,731] |
|  | 5 years, discounting rate = 4% | 94,913 [94,794; 95,039] | 4.561 | 282,393,955 [281,501,382; 283,236,792] |
|  | 10 years, undiscounted | 200,952 [200,567; 201,332] | 9.657 | 597,647,864 [595,680,177; 599,670,245] |
|  | 10 years, discounting rate = 3% | 176,895 [176,570; 177,216] | 8.501 | 526,122,041 [524,425,262; 527,864,155] |
|  | 10 years, discounting rate = 4% | 169,938 [169,628; 170,241] | 8.167 | 505,435,208 [503,808,818; 507,108,274] |
| No excess mortality | 1 year, undiscounted | 20,754 [20,742; 20,766] | 0.997 | 61,764,712 [61,583,026; 61,939,174] |
|  | 5 years, undiscounted | 102,563 [102,435; 102,697] | 4.929 | 305,153,172 [304,191,712; 306,068,575] |
|  | 5 years, discounting rate = 3% | 96,795 [96,677; 96,920] | 4.652 | 287,993,150 [287,087,745; 288,859,116] |
|  | 5 years, discounting rate = 4% | 95,017 [94,902; 95,139] | 4.566 | 282,702,955 [281,814,863; 283,553,654] |
|  | 10 years, undiscounted | 201,443 [201,081; 201,814] | 9.681 | 599,124,738 [597,215,862; 601,090,358] |
|  | 10 years, discounting rate = 3% | 177,305 [177,002; 177,622] | 8.521 | 527,352,287 [525,688,067; 529,061,844] |
|  | 10 years, discounting rate = 4% | 170,324 [170,038; 170,629] | 8.186 | 506,595,341 [504,995,907; 508,230,770] |

\*8% excess mortality the 1<sup>st</sup> year, (8% x 8/9) the 2<sup>nd</sup> year, (8% x 7/9) the 3<sup>rd</sup> year, ... (8% x 1/9) the 9<sup>th</sup> year, and 0% the 10<sup>th</sup> year.
\*\*Numbers shown in this cell and cells below are the mean [95% confidence interval], based on 1000 simulations replicating the cohort.

**Table 3.** Prediction of the total ART costs spared (€) hypothesizing that 50% of the persons living in France and fulfilling QUATUOR inclusion criteria (n = 0.5 × 104,042 = 52,021) would switch from the 7/7-days strategy to the 4/7-days strategy.

| **Excess mortality in the target population as compared to the mortality in the whole French population** | **Time Horizon and discounting rate** | **Life-years expected, totaled for the target population adopting the 4/7-days strategy ($n = 52,021$)** | **ART costs (€) expected to be spared with the 4/7-days strategy as compared to the 7/7-days strategy, totaled for the whole target population ($n = 52,021$)** |
| --- | --- | --- | --- |
| Linear decreasing of excess mortality each year (from 8% to 0%)* | 1 year, undiscounted | 51,875 [51,855; 51,893]** | 154,376,338 [154,084,190; 154,659,273]** |
|  | 5 years, undiscounted | 256,166 [255,950; 256,373] | 762,119,690 [760,574,914; 763,689,577] |
|  | 5 years, discounting rate = 3% | 241,764 [241,565; 241,955] | 719,278,520 [717,825,049; 720,755,214] |
|  | 5 years, discounting rate = 4% | 237,324 [237,130; 237,510] | 706,071,091 [704,644,776; 707,519,047] |
|  | 10 years, undiscounted | 502,838 [502,232; 503,438] | 1,495,428,616 [1,492,194,706; 1,498,663,767] |
|  | 10 years, discounting rate = 3% | 442,609 [442,108; 443,117] | 1,316,360,527 [1,313,562,448; 1,319,150,447] |
|  | 10 years, discounting rate = 4% | 425,191 [424,720; 425,674] | 1,264,572,236 [1,261,897,374; 1,267,262,876] |
| Fixed 8% excess mortality | 1 year, undiscounted | 51,875 [51,855; 51,893] | 154,376,338 [154,084,190; 154,659,273] |
|  | 5 years, undiscounted | 256,124 [255,911; 256,329] | 761,997,016 [760,455,377; 763,546,893] |
|  | 5 years, discounting rate = 3% | 241,726 [241,530; 241,916] | 719,167,435 [717,714,375; 720,626,737] |
|  | 5 years, discounting rate = 4% | 237,287 [237,097; 237,473] | 705,963,538 [704,539,672; 707,396,407] |
|  | 10 years, undiscounted | 502,391 [501,773; 503,019] | 1,494,095,774 [1,490,830,412; 1,497,353,746] |
|  | 10 years, discounting rate = 3% | 442,248 [441,731; 442,762] | 1,315,282,123 [1,312,443,479; 1,318,114,902] |
|  | 10 years, discounting rate = 4% | 424,854 [424,369; 425,340] | 1,263,565,359 [1,260,851,140; 1,266,296,360] |
| No excess mortality | 1 year, undiscounted | 51,886 [51,867; 51,903] | 154,406,548 [154,123,579; 154,690,755] |
|  | 5 years, undiscounted | 256,411 [256,206; 256,613] | 762,855,498 [761,310,640; 764,403,600] |
|  | 5 years, discounting rate = 3% | 241,990 [241,799; 242,179] | 719,957,232 [718,504,506; 721,419,155] |
|  | 5 years, discounting rate = 4% | 237,544 [237,358; 237,728] | 706,732,292 [705,307,916; 708,167,615] |
|  | 10 years, undiscounted | 503,619 [503,024; 504,209] | 1,497,784,519 [1,494,587,347; 1,501,089,883] |
|  | 10 years, discounting rate = 3% | 443,271 [442,781; 443,765] | 1,318,354,725 [1,315,609,783; 1,321,198,345] |
|  | 10 years, discounting rate = 4% | 425,818 [425,353; 426,283] | 1,266,462,801 [1,263,840,805; 1,269,173,228] |

\*8% excess mortality the 1<sup>st</sup> year, (8% x 8/9) the 2<sup>nd</sup> year, (8% x 7/9) the 3<sup>rd</sup> year, ... (8% x 1/9) the 9<sup>th</sup> year, and 0% the 10<sup>th</sup> year.
\*\*Numbers shown in this cell and cells below are the mean [95% confidence interval], based on 1000 simulations replicating the cohort.

## DISCUSSION

This study is the first to date reporting in details cost-effectiveness aspects comparing a triple ART taken 7 days a week with a triple ART taken 4 consecutive days on and 3 days off. The main analysis indicated that the relevant differences between the two compared strategies only concern ART costs (Table 1). Adopting a 4/7-days strategy would spare 40% of the ART costs, and the latter costs were estimated to represent about 80% of the direct medical costs, yielding a sparing of approximately 1/3 of the total medical costs. Hypothesizing the adoption of the 4/7-days strategy in 20% of the persons eligible–a conservative assumption, corresponding annual savings of M€ 62 would be expected at the national French level (Table 2).

The costs reported in this study for the standard of care (7/7-days strategy) are in agreement with national costs in France reported by other sources. For example, Rachas et al reported an amount of € 1.22 billions spent in 2019 by the French social security for the total medical resources of the 151,346 PLWH,^21^ leading to € 8,060 mean costs per person, a value close to that of € 8,089 issued from the present analysis of the QUATUOR trial (Table 1). In addition, open access data^22^ further detail these total expenses, and report medication costs totalizing € 1,014,274,924 in 149,090 persons, these medication costs representing 83% of the total medical costs, and leading to € 6,803 mean costs per person, a value close to that of € 6,929 issued from the present analysis of the QUATUOR trial (see individual costs for ART + co-medications in Table 1).

We also explored treatment success instead of noninferiority as the effectiveness criterion in cost-effectiveness analyses, and the corresponding results provide further critical information on the interest of the 4/7-days strategy in terms of cost-effectiveness issues. First, the simulations indicate that the proportions of treatment success observed in the two arms of the QUATUOR trial were not significantly different (see results section). Second, even when considering the higher proportion of successes observed in the 7/7-days strategy, the simulations replicating the QUATUOR trial indicate that obtaining an additional treatment success in the 7/7-days strategy as compared to the 4/7-days strategy requires very substantial additional costs, with a corresponding estimated ICER of € 220,383 per additional treatment success. Furthermore, acceptability curves indicate that only 15% of the simulations resulted in an ICER ≤ 100 K€, and only 23% of the simulations resulted in an ICER ≤ 3 gross domestic product per inhabitant (Fig. 2b). It is worth mentioning that the threshold of 3 gross domestic product per inhabitant has been proposed for cost-utility analyses in which costs are contrasted to quality adjusted life years, and it is very unlikely that a given person switching to the 4/7-days might experience a decrease as important as 1 quality-adjusted life year. Indeed, any treatment failure in persons switching from the 7/7-days to the 4/7-days is likely expected to be detected rapidly, leading to prompt and appropriate treatment modifications. Therefore, only modest unfavorable consequences should be observed in the rare individuals for which the 4/7-days strategy would appear detrimental. Moreover, reinforcing the surveillance of treatment success in persons under the 4/7-days strategy could be proposed (and for example, especially during the 2 years following the switch date), and even assuming very frequent controls, or hypothesizing additional tests (e.g., potential prescriptions of HIV DNA genotyping for comforting decision-making of the switch towards the 4/7-days strategy) the relative corresponding additional costs would still lead to a balance favoring very substantial economic savings with the 4/7-days strategy.

The 4/7-days strategy addresses concerns about the burden of daily medications in PLWH. Addressing similar concerns, the interest of long-term injections has attracted a great deal of attention, especially therapies based on dual injections of cabotegravir and rilpivirine (about 6 per year).

However, as mentioned by Nachege et al, additional data for better documenting the cost-effectiveness of such therapies in various settings are needed.^23^ The present study framework and results might be considered as a useful reference for future studies in the field. For example, considering the current official costs of € 7082 for 6 dual injections of cabotegravir and rilpivirine in France,^24^ and hypothesizing that the remaining medical costs and health outcomes observed in persons treated with such dual injections would be those observed in the present study, a cost-effectiveness analysis would categorize such long-term injections as dominated by the 4/7-days investigated here.

This study has some limitations. ART costs were estimated considering nongeneric medications. However, in France, the most frequent therapies are currently based on drugs which are not commercialized under nongeneric licenses. Cost valuation of laboratory tests in this study are somehow over-estimated for two main reasons. First, the persons of the trial were inherently more strictly controlled than they would be otherwise. Second, in practice, some PLWH may choose to take advantage of daily-hospital stays allowed by the French system for controlling their health, and fees for such stays include some laboratory tests while all laboratory tests of the study were valued as if they were independently prescribed. Nevertheless, as above mentioned, savings with the 4/7-days strategy would remain very substantial assuming more frequent laboratory tests. Study limitations also concern generalization issues. Cost estimates presented here cannot be extended to other countries than France. In particular, ART costs are much lower in low- and middle-income countries. Therefore, in corresponding countries, adopting a 4/7-days strategy would likely result in more modest proportional savings, considering the lower weight of ART in the total medical costs of HIV management. Future investigations on this topic constitute attractive research perspectives.

In conclusion, this cost-effectiveness study estimates that switching from a 7/7-days to a 4/7-days strategy enables savings amounting to 1/3 of the total direct medical costs while maintaining ART effectiveness, in PLWH in France, with a triple ART and a controlled viral load. The study favors generalizing the adoption of the 4/7-days strategy in France.

## DATA AVAILABILITY

Data requests can be submitted to the study steering committee of the present study (by email to) and must be approved by the French data protection authority (la Commission Nationale de l’Informatique et des Libertés [CNIL]). According to French law, anyone wishing an access to cohort or clinical study data on humans is required to ask permission to the French data protection authority (the CNIL). The permission process is initiated by completing a form that can be provided by Lambert Assoumou. For further information, please see https://www.cnil.fr/.

The study steering committee will evaluate each proposal for compatibility with general objectives, ethical approval, informed consent forms of the ANRS 170 QUATUOR project, and for potential overlap with ongoing work.

## ACKNOWLEDGEMENTS

We thank the participants of the ANRS 170 QUATUOR study for their time and dedication to this research for the benefit of their community. We thank the monitoring and data management staff who made this study possible through continuous interactions with study staff at study sites and between visits. We also thank the staff at the study sites, without whom this study would not have been possible.

## AUTHOR CONTRIBUTIONS

L.A. and G.H initiated the study; L.A. and G.H. supervised the study; G.H. designed the experimental plan; S.D. collected cost data and managed study data; S.D., G.H., and P.M. performed the analyses; S.D. and L.A. can take responsibility for the integrity of the data, and G.H. for the accuracy of data analyses; G.H. prepared the first draft of the manuscript, with the help of L.A. and P.M.; all authors (C.A., J-C.A., K.A., L.A., J.C., D.C., S.D, J.G., S.G., G.H., C.K., K.L., S.L-N., R.L., L.M-J., P.M., D.P., and P.D.T.) contributed to interpretation of the data, critically revised the manuscript, and approved the final version.

